# Meta-Analysis and Structural Dynamics of the Emergence of Genetic Variants of SARS-CoV-2

**DOI:** 10.1101/2021.03.06.21252994

**Authors:** Nicolas Castonguay, Wandong Zhang, Marc-Andre Langlois

## Abstract

The novel Severe Acute Respiratory Syndrome Coronavirus 2 (SARS-CoV-2) emerged in late December 2019 in Wuhan, China, and is the causative agent for the worldwide COVID-19 pandemic. SARS-CoV-2 is a positive-sense single-stranded RNA virus belonging to the betacoronavirus genus. Due to the error-prone nature of the viral RNA-dependent polymerase complex, coronaviruses are known to acquire new mutations at each cycle of genome replication. This constitutes one of the main factors driving the evolution of its relatively large genome and the emergence of new genetic variants. In the past few months, the identification of new B.1.1.7 (UK), B.1.351 (South Africa) and P.1 (Brazil) variants of concern (VOC) have highlighted the importance of tracking the emergence of mutations in the SARS-CoV-2 genome that impact transmissibility, virulence, and immune and neutralizing antibody escape. Here we analyzed the appearance and prevalence trajectory over time of mutations that appeared in all SARS-CoV-2 genes from December, 2019 to April, 2021. The goal of the study was to identify which genetic modifications are the most frequent and study the dynamics of their propagation, their incorporation into the consensus sequence, and their impact on virus biology. We also analyzed the structural properties of the spike glycoprotein of the B.1.1.7, B.1.351 and P.1 variants for its binding to the host receptor ACE2. This study offers an integrative view of the emergence, disappearance, and consensus sequence integration of successful mutations that constitute new SARS-CoV-2 variants and their impact on neutralizing antibody therapeutics and vaccines.

**IMPORTANCE:** SARS-CoV-2 is the etiological agent of COVID-19, which has caused > 3.4 million deaths worldwide as of April, 2021. Mutations occur in the genome of SARS-CoV-2 during viral replication and affect viral infectivity, transmissibility, and virulence. In early March 2020, the D614G mutation in the spike protein emerged, which increased viral transmissibility and is now found in over 90% of all SARS-CoV-2 genomic sequences in GISAID database. Between October and December 2020, B.1.1.7 (UK), B.1.351 (South Africa) and P.1 (Brazil) variants of concern (VOCs) emerged, which have increased neutralizing antibody escape capabilities because of mutations in the receptor binding domain of the spike protein. Characterizing mutations in these variants is crucial because of their effect on adaptive immune responses, neutralizing antibody therapy, and their impact on vaccine efficacy. Here we tracked and analyzed mutations in SARS-CoV-2 genes since the beginning of the pandemic and investigated their functional impact on the spike of these three VOCs.

**Graphical Abstract:** 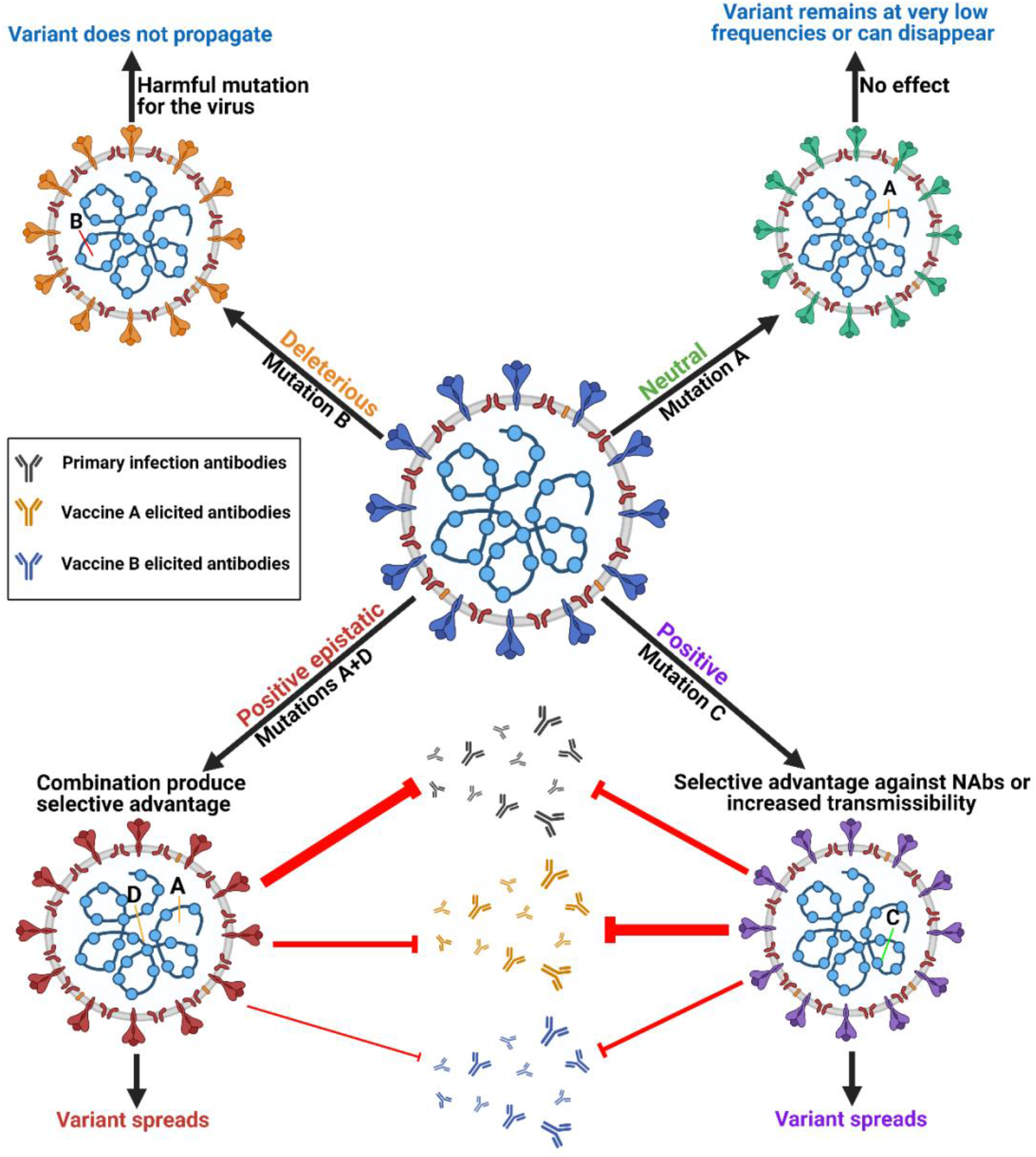

## INTRODUCTION

In late December 2019, a new betacoronavirus known as Severe Acute Respiratory Syndrome Coronavirus 2 (SARS-COV-2) emerged in the city of Wuhan in the province of Hubei, China (1). SARS-CoV-2 is the etiological viral agent for the worldwide COVID-19 pandemic resulting in more than 162 million infected and 3.4 million deaths worldwide as of April, 2021 (2,3). SARS-CoV-2 is an enveloped, positive-sense single-stranded RNA (+ssRNA) virus with a genome length of 29,811 nucleotides (4). The mutation rates of RNA viruses are generally higher than that of DNA viruses because of the low fidelity of their viral RNA polymerases (5,6). Mutations occur when viral replication enzymes introduce errors in the viral genome resulting in the creation of premature termination codons, deletions and insertions of nucleotides that can alter open reading frames and result in amino acid substitutions in viral proteins. These mutations combined with the selective pressure of the human immune system lead to the selection and evolution of viral genomes (6,7). However, coronaviruses are one of the few members of the RNA virus family that possess limited but measurable proofreading ability via the 3’ -to 5’-exoribonuclease activity of the non-structural viral protein 14 (nsp14) (8,9). Coronaviruses are therefore expected to evolve through genetic drift much slower than other RNA viruses that do not have this ability, such as influenza viruses (8,10). Additionally, SARS-CoV-2 and other coronaviruses have low known occurrences of recombination between family members (i.e., genetic shift), and therefore are mostly susceptible to genetic drift (11).

SARS-CoV-2 has reached pandemic status due to its presence on every continent and has since maintained a high level of transmissibility across hosts of various ethnical and genetic backgrounds (2, 12). Moreover, SARS-CoV-2 infections have been reported to naturally infect minks, ferrets, cats, tiger, and dogs, which allows the virus to replicate in completely new hosts and mutate to produce new variants and possibly new strains (13,14). In March 2020, the now dominant D614G mutation first emerged in the spike protein (S) of SARS-CoV-2. The S protein is present as a trimer at the surface of the viral envelope and is responsible for attachment of the virus to the human angiotensin converting enzyme 2 (hACE2), the entry receptor for SARS-CoV-2 into human cells (15). Published evidence has now shown that D614G increases viral fitness, transmissibility and viral load but does not directly affect COVID-19 pathogenicity (16,17,18,19). Additionally, emerging evidence indicates that D614G may have epistatic interactions that exacerbate the impact of several other independent mutations (19). Mutations in the S protein, and particularly in the receptor binding domain (RBD), are of very high concern given that they can directly influence viral infectivity, transmissibility, and resistance to neutralizing antibodies and T cell responses.

New mutations are frequently and regularly detected in the genome of SARS-CoV-2 through whole genome sequencing; however, very few of these mutations make it into the transmitted viral consensus sequence. The reference strain is generally regarded as the dominant transmitted strain at a given time. Its sequence is determined by aligning large numbers of recently sequenced genomes and establishing a consensus sequence composed of the highest frequency nucleotide for each position in the viral genome. A genetic variant is a version of the reference strain that has acquire one or several new mutations and acts as the founder for further genetic diversification and evolution. Mutations arise regularly in the reference strain, but few are longitudinally conserved. Genetic variants are therefore the rare successful offshoots of the reference strain.

Some variants rise rapidly in frequency and then collapse and disappear, while others will rise and overtake the frequency of the reference strain and become the new reference. There are three main genetic variants that have emerged in the past few months with sustained upward frequency trajectories. The first is the UK variant, also known as B.1.1.7/501Y.V1 (B1.1.7). It was first detected in September 2020 and is now present worldwide and poised to become the new reference strain (20,21). The South African variant, also known as B.1.351/501Y.V2 (B.1.351), was first reported in October 2020 and is now increasing in prevalence in South Africa, Europe, and North America (21,22). The Brazilian variant, P.1/501Y.V3 (P.1), was first detected in travelers from Brazil that landed in Japan in January 2021 (23). It has since been identified in 42% of specimens in the Amazonian city of Manaus and in the U.S. at the end of January 2021 (23). These three variants are associated with increased resistance to neutralizing antibodies (Nabs) and all possess the N501Y mutation, which is a mutation in the RBD that is critical for the spike protein to interact with hACE2 (24,25,26). This mutation is reported to cause increased resistance to Nabs, increased transmissibility, and increased virulence in animal models (27). In addition to the N501Y mutation, both the South African and Brazil variants possesses RBD mutations K417N(T) and E484K, which are also associated with further increased Nabs escape capabilities (24,25).

Here we present a retrospective metadata analysis of mutations throughout the SARS-CoV-2 genome that reached at least a 1% worldwide frequency between December 2019 and January 2021. We specifically investigated their frequency trajectory over time and their fixation into the reference sequencing using the Global Initiative on Sharing Avian Influenza Data (GISAID) (28). Additionally, we analyzed mutations in the S protein of the B.1.1.7, B.1.351 and P.1 variants and illustrated their impact on molecular interactions between the S protein and hACE2 and their potential impact on Nabs.

## MATERIALS AND METHODS

### Data collection and mutational analysis

Genomes uploaded to the GISAID EpiCoV™ server database were analyzed from December 1^st^, 2019, to December 31^st^, 2020, and selected viral sequences with submission dates from December 1^st^, 2019 to January 6, 2021. We first selected recurring mutations that were present in more than 500 reported genomes by August 2020, and another selection was made in January 2021 to capture recurring mutations present in more than 4000 reported genomes in GISAID. This strategy allowed us to study mutations reaching at least 1% in worldwide frequency. We filtered through 309,962 genomes for the analysis of selected mutations.

The worldwide frequency of the hCoV-19/Wuhan strain, and hCoV-19/D614G, B.1.1.7, B.1.351, and P.1 variants were analyzed from December 1^st^, 2019 to April 30^th^ 2021. For the analysis of the mutations in B.1.1.7, B.1.351, and P.1 variants, we used the GISAID EpiCoV™ server database. Viral sequences on GISAID with submission dates between December 1^st^, 2019 and April 30^th^ 2021 were selected for the analysis. We filtered through 1,090,689 genomes for the analysis of the variants. Only complete SARS-CoV-2 genomes (28 to 30 Kbps) isolated from human hosts were analyzed. MUSCLE alignment tool on UGene, and SnapGene was used to determine the nucleotide mutations and codon changes of the non-synonymous and synonymous mutations by sequence alignments to the NCBI SARS-CoV-2 reference genome (NC_045512). All genomes uploaded to the GISAID database that were used in this study are presented in **Supplementary Table 1**. Graphs of mutations and variants were performed in RStudio with timelines, and genomes illustrations were produced in Biorender.

### Structural modeling

Mutations in the spike protein in complex with hACE2 were analyzed using a mutagenesis tool for PyMOL (PDB: 7A94). Visualizations of mutations in the B.1.1.7, B.1.351 and P.1 variants were produced using the spike protein closed conformation (PDB: 6ZGE), interaction with hACE2 (PDB: 7A94), interaction with C102 Nab (PDB: 7K8M), and interaction with C121 Nab (PDB: 7K8X). Figures and rendering were prepared with PyMOL.

## RESULTS

### Identification of emerging mutations in various SARS-CoV-2 genes

Emerging mutations in the SARS-CoV-2 genome were investigated to illustrate the fluctuations of these mutations during a period of twelve months. We tracked genome mutations with a worldwide frequency greater than 1% from December 2019 to December 31st, 2020 in the GISAID database. Genes NSP8, NSP10, NS6, NS7a, and E are not illustrated in **Figure 1** given that they did not display mutations with frequencies sufficiently high to meet our inclusion criteria during the study period. This is indicative that these are some of the most conserved sequences of the SARS-CoV-2 genome. Viral proteins that have low mutation frequencies could be due to evolutionary constraints. Some of these viral proteins may be evolutionarily conserved and play important roles in virus assembly and stability, genome replication, and viral release from infected cells. Our analysis highlights the fixation of the D614G mutation in the S protein and that of the P323L mutation in the RNA-dependent RNA polymerase (RdRp) (**Fig.1**). These are the only mutations to have successfully become part of the reference sequence as of December 2020. They appeared to have emerged simultaneously in January, 2020 and became present in more than 90% of all sequenced genomes by June, 2020. Additionally, some other mutations also emerged rapidly but then stabilized in frequency or faded out. For example, Q57H (NS3), R203K (N), G204R (N) are mutations that emerged rapidly and appeared to have stabilized at a frequency of 15% to 40%. Others like I120F (NSP2), L37F (NSP6), S477N (S), and L84S (NS8) illustrate mutations that emerged rapidly and then faded-out just as quickly. We also demonstrate that most genes in SARS-CoV-2 have mutations with overall frequencies lower than 10% (**Fig.1)**. These mutations are also summarized into **Table 1**, which illustrates nucleotide substitution producing the amino acid change, the frequency, and their respective effects. Globally, there is an uneven effort to test for SARS-CoV-2 infections in the population and sequence the viral genomes in those infected. As a result, emerging mutations go unreported until they reach countries with more intensive sequencing capabilities. Therefore, mutations presented here vastly underrepresent the global landscape of mutation frequency dynamics.

**Figure 1:**
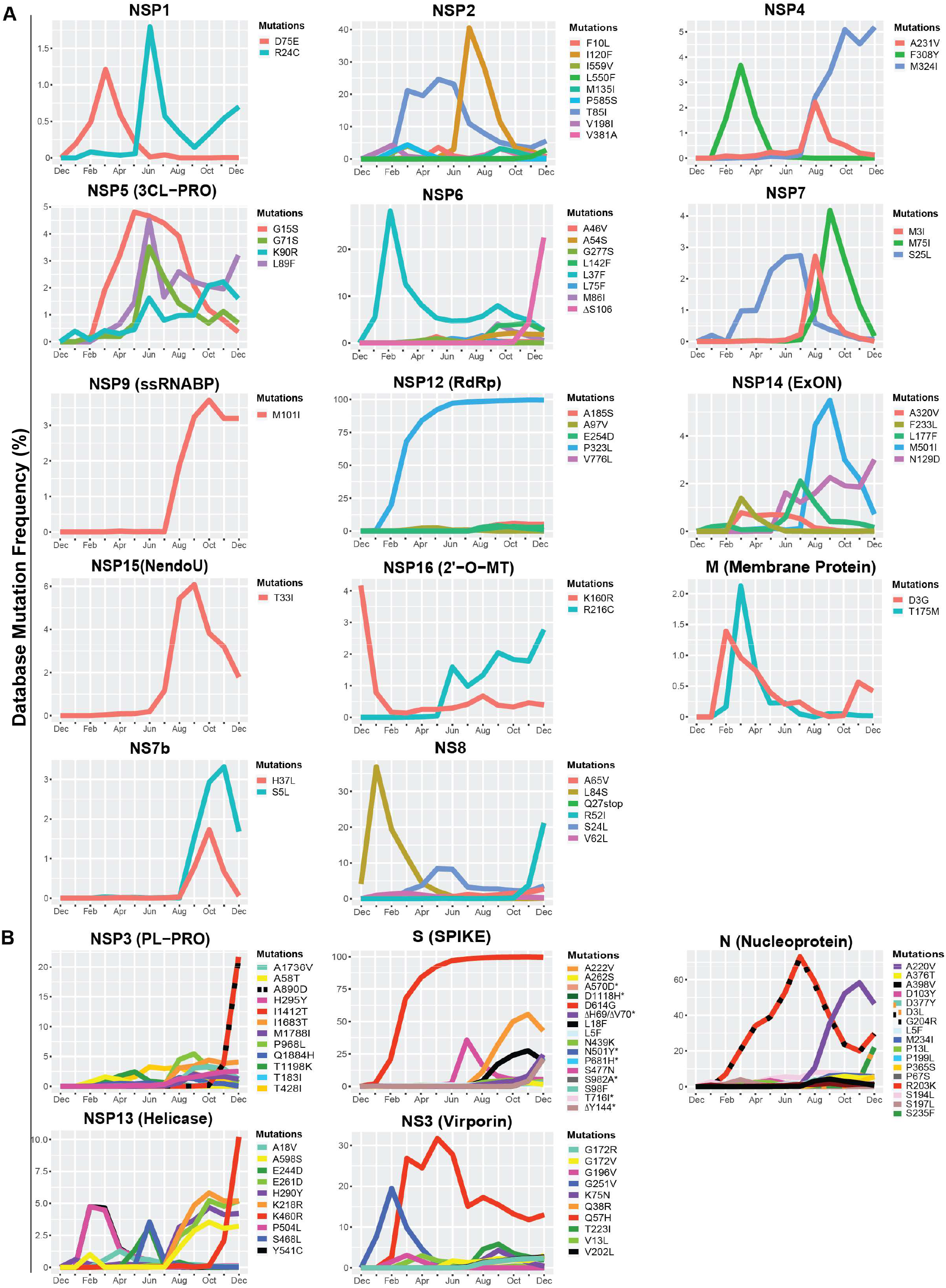
Variations in mutations and mutation frequencies in SARS-CoV-2 genes. The occurrence and frequency of mutations in various SARS-CoV-2 genes are presented for the period between December 2019 to January 1^st^, 2021. Plotted are mutations that reached at least a 1% worldwide frequency. SARS-CoV-2 genes are represented with the function of the genes in parentheses. A) Genes with <10 mutations. B) Genes with >10 mutations. Graphs were generated using RStudio. *overlapping curves

**Table 1:**
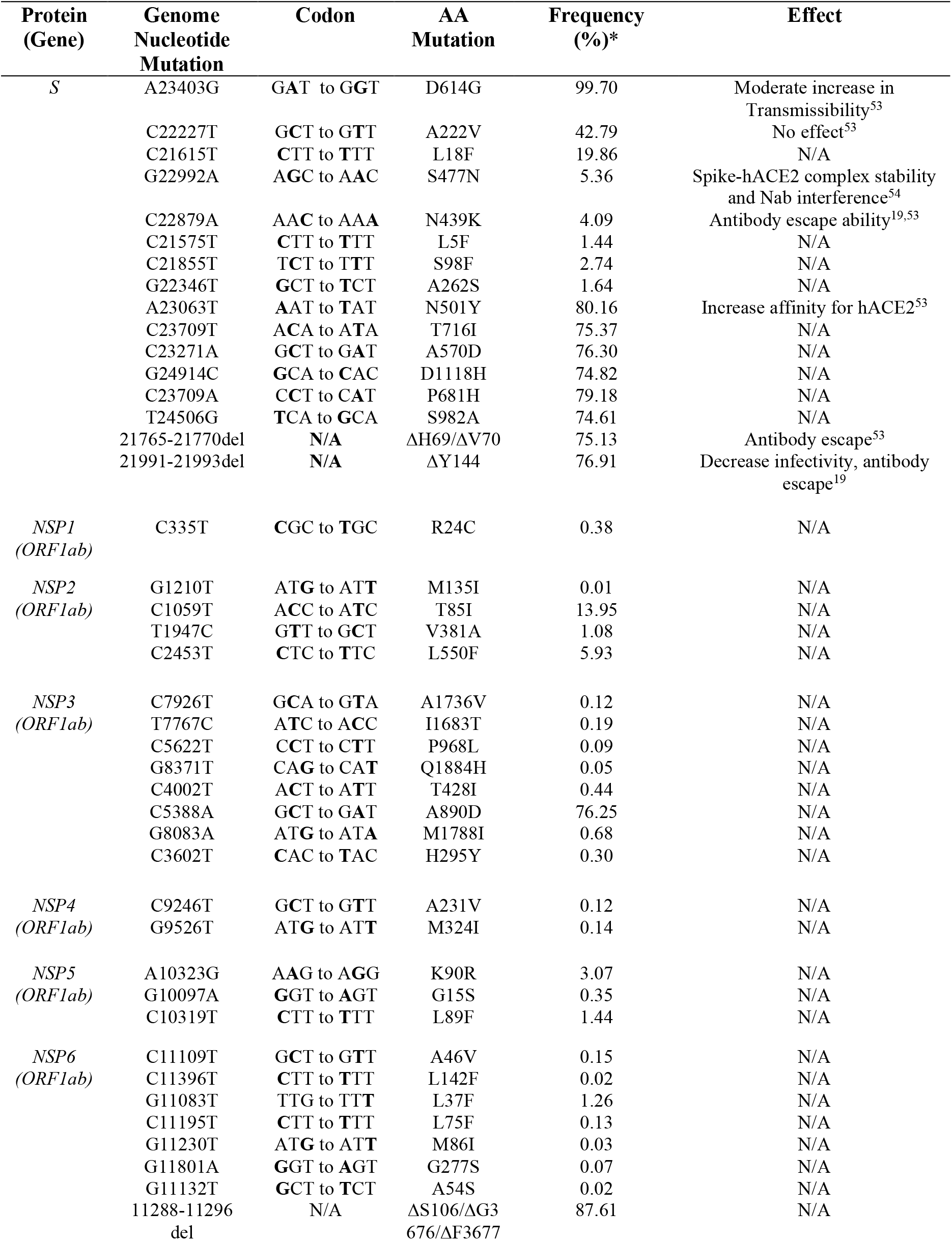
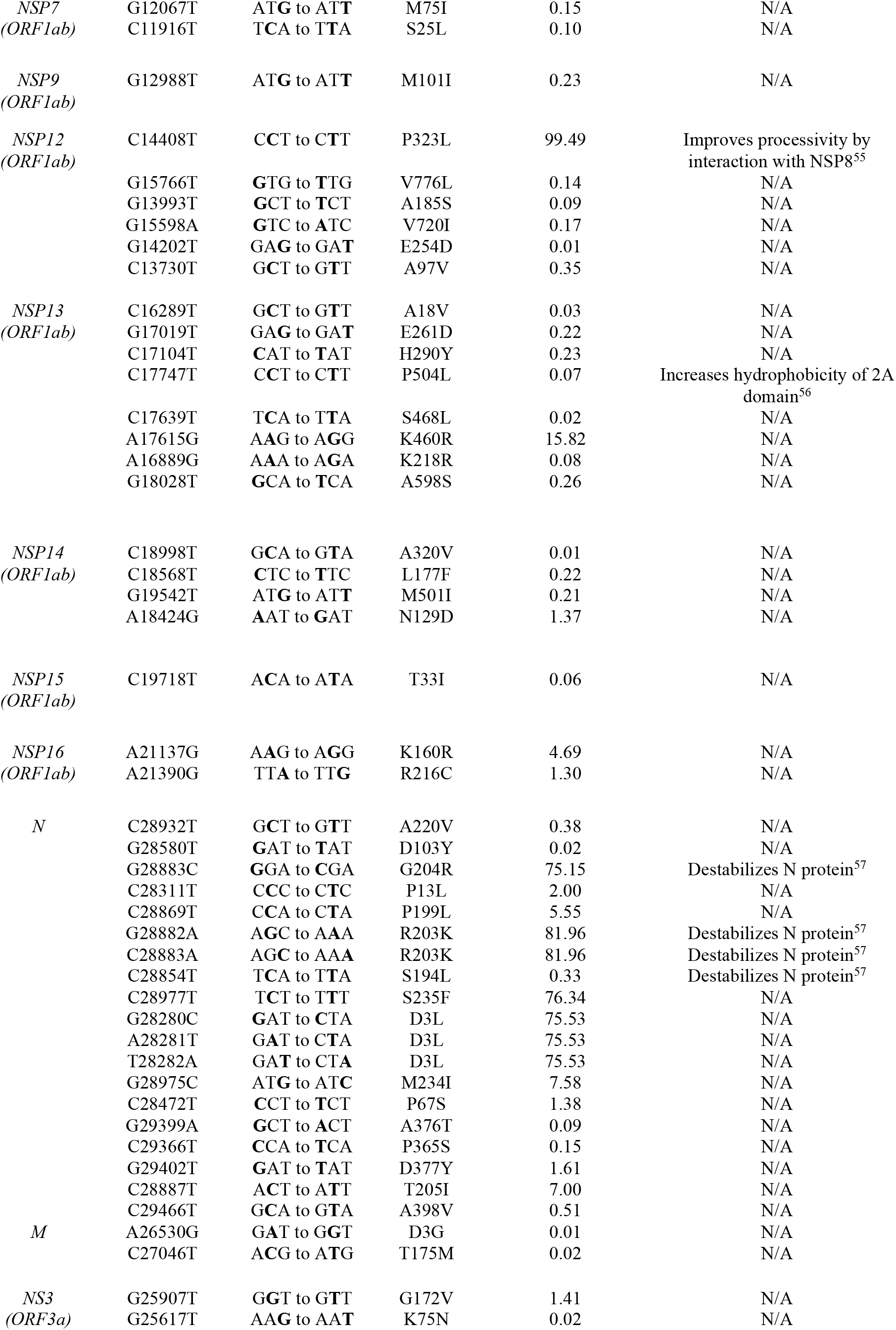
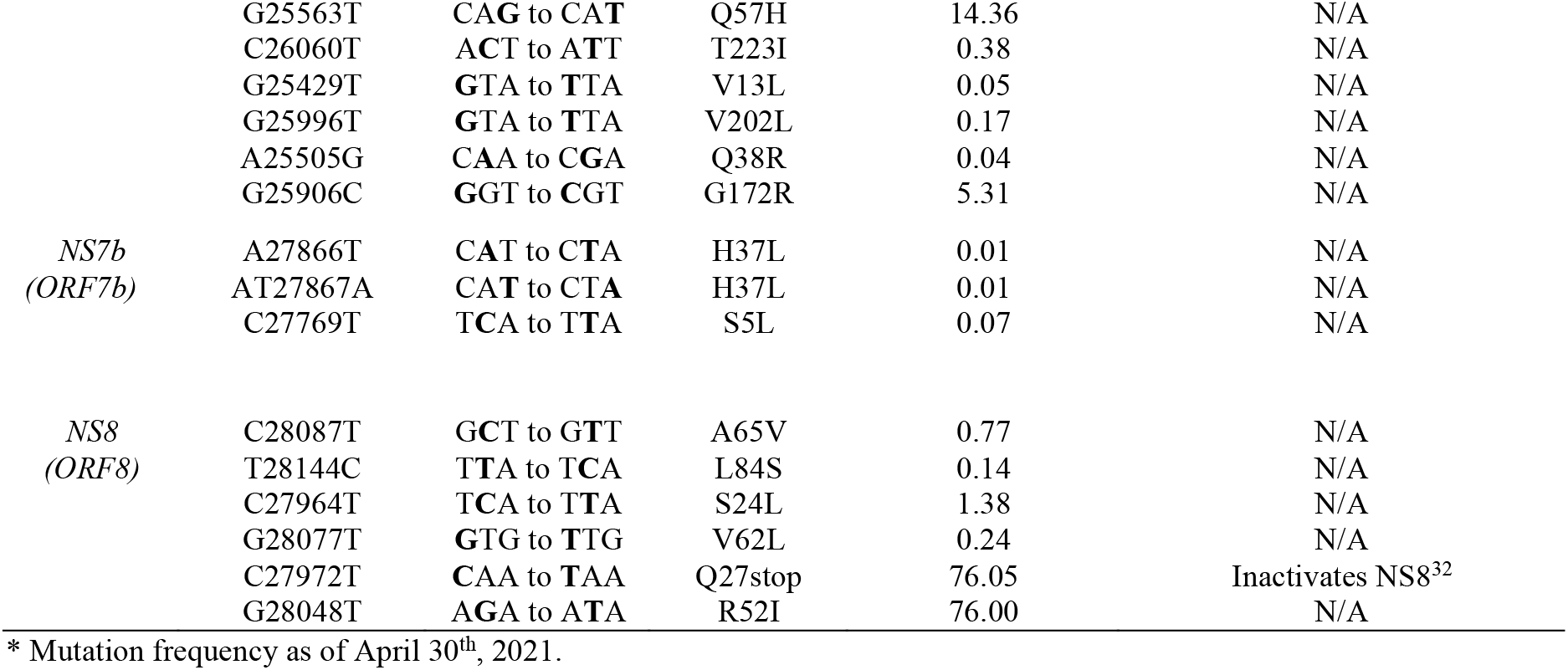
Non-synonymous mutations in SARS-CoV-2 genes with a worldwide frequency ≥0.01%.

### Geographic localization and timeline of the viral genes with mutations higher than 50% frequency

We next turned our attention to viral genes where at least one mutation reached a frequency higher than 50%. Only three genes met this criterion: NSP12, S, and N. We then took their mutation graphs and added worldwide geographic data provided from GISAID. Geomaps are useful to determine if a mutation is a localized and regional event or is found worldwide. In the S protein, D614G is found worldwide with initially higher reported cases in the US, UK, and Australia, probably due to more intense large-scale testing and sequencing, while A222V is mostly reported in the UK, but has not yet been reported in South America, Central and East African regions (**Fig. 2A)**. Like D614G, P323L in the RdRp is also found worldwide, with higher reported cases in the US, UK, and Australia (**Fig. 2B)**. The N gene does not yet have successful mutations that have attained reference sequence status, but R203K, G204R, A220V, D3L, and S235F all reached a frequency of 50% or higher during the study period. A220V emerged in August of 2020 and reached a frequency higher than 50% in October of 2020 (**Fig. 2D**). Mutations G204R, R203K and A220V in N gene are reported at high frequency in the UK, but were not been detected in South America, Central, East, and South African regions (**Fig. 2C)**. We also observe the emergence of several mutations in S and N present in the B.1.1.7 variant, which now have a frequency higher than 65% (**Fig. 2C, 2D and Table 2**). The analysis of these data illustrate the localization of the most prevalent mutations to date, which appear to be mostly present in Western high-income countries. This, however, is undoubtable attributable to overall more intense testing and sequencing.

**Figure 2:**
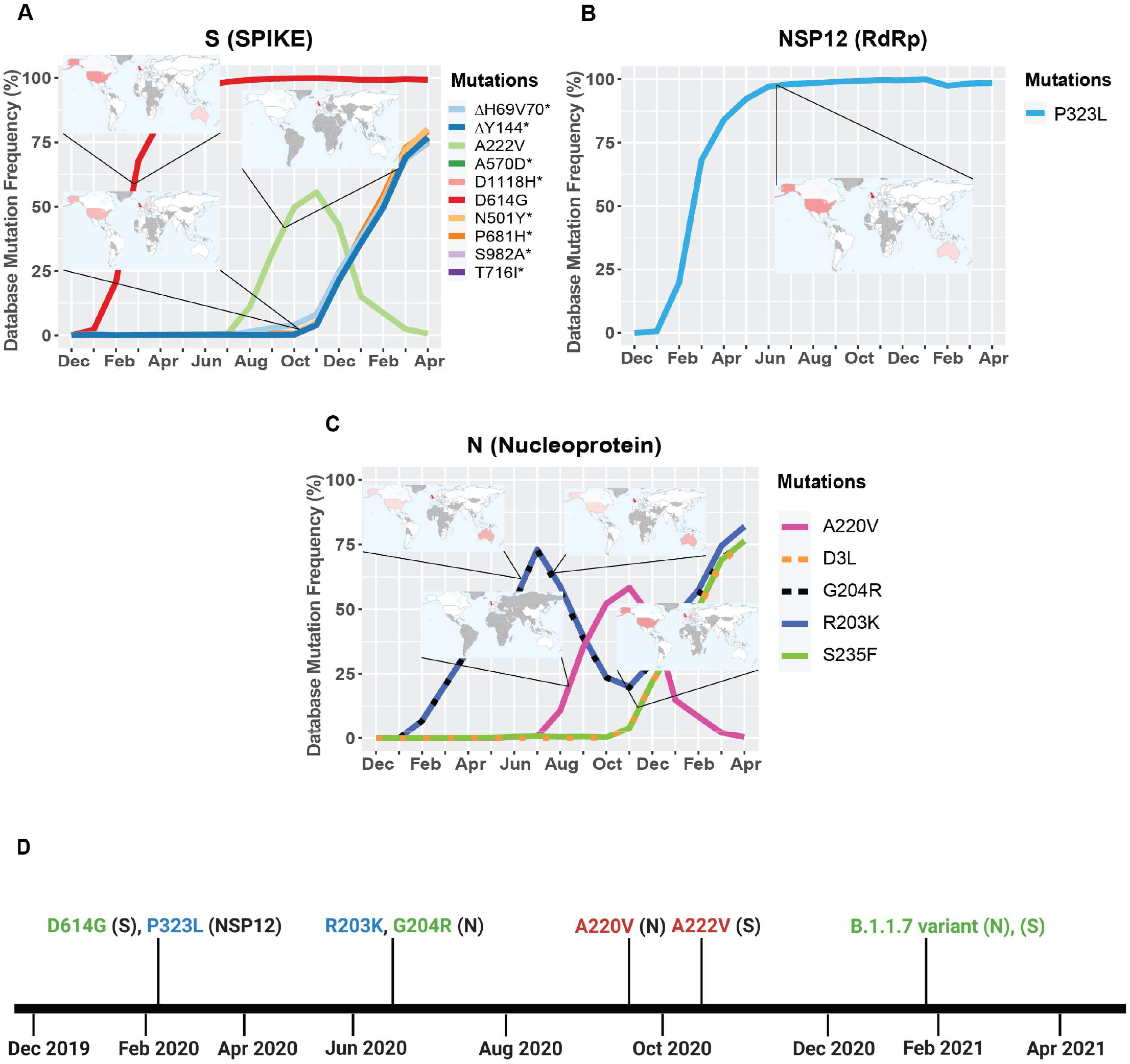
Geographic location and timeline of dominant mutations in NSP12, S, and N genes. A) Frequency of S protein mutations with corresponding geographic maps. B) Frequency of RdRp mutations with corresponding geographic maps. C) Frequency of Nucleoprotein mutations with corresponding geographic maps. D) Timeline of the appearance of mutations reaching a frequency higher than 50% worldwide between December, 2019 and April, 2021. For the geomaps: a low frequency of reported cases of the mutations is represented in white, while higher frequencies are represented from pink to red, and grey represents no data. All maps were taken from GISAID. Graphs were generated using RStudio and Biorender. *overlapping curves

**Table 2:**
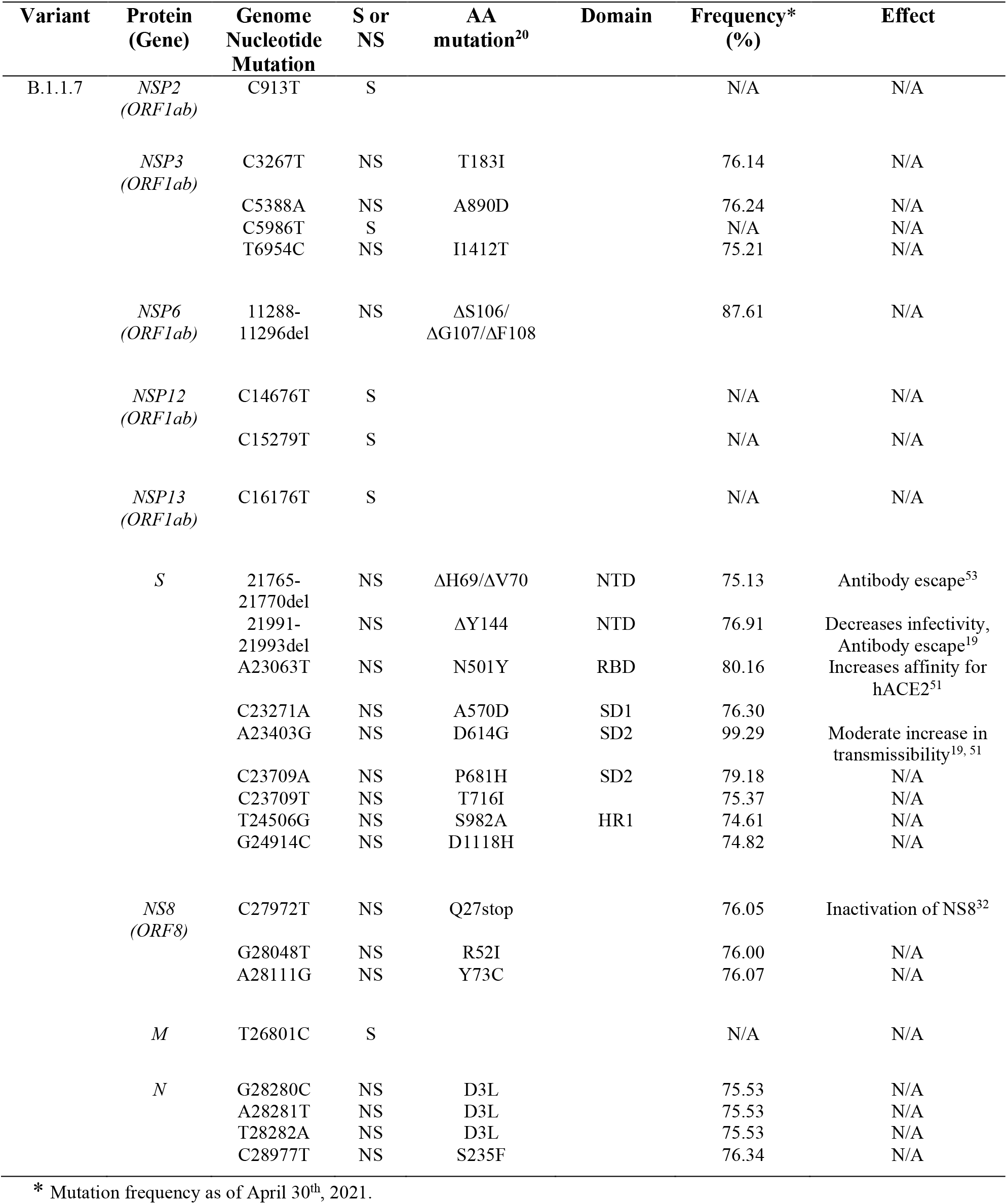
Synonymous, non-synonymous and deletion mutations in the B.1.1.7 variant

### Localization and molecular interactions of prevalent S protein mutations

Here we illustrate the molecular interactions and spatial localization of the mutations in the S protein. PyMOL was used to model structures of the S protein binding to ACE2 and we analyze the possible effects of specific mutations at given positions in the protein. **Figure 3A** presents an overview of the physical positions of S protein mutations. The A222V mutation has no reported effects on protein stability, neutralizing antibody escape, and affinity for hACE2 **(Table 1)**. The substitution from A to V results in a low steric clash between neighboring residues (**Fig. 3B)**. The S477N substitution in the RBD enables increased stability during hACE2-RBD interactions (**Fig. 3C, Table 1)**. In the N-terminal domain (NTD), mutation L18F leads to a steric clash between neighboring residues (**Fig. 3D)**. However, this does not appear to impact the stability of the protein given that no such effects have been reported so far (**Table 1)**. In the closed conformation, D614 makes an ionic bond with K854 in the S2 subunit of another S protein monomer (**Fig. 3F)** (29). In the open conformation, D614 (or G614) doesn’t make interactions or display steric clashes with neighboring residues (**Fig. 3E)**.

**Figure 3:**
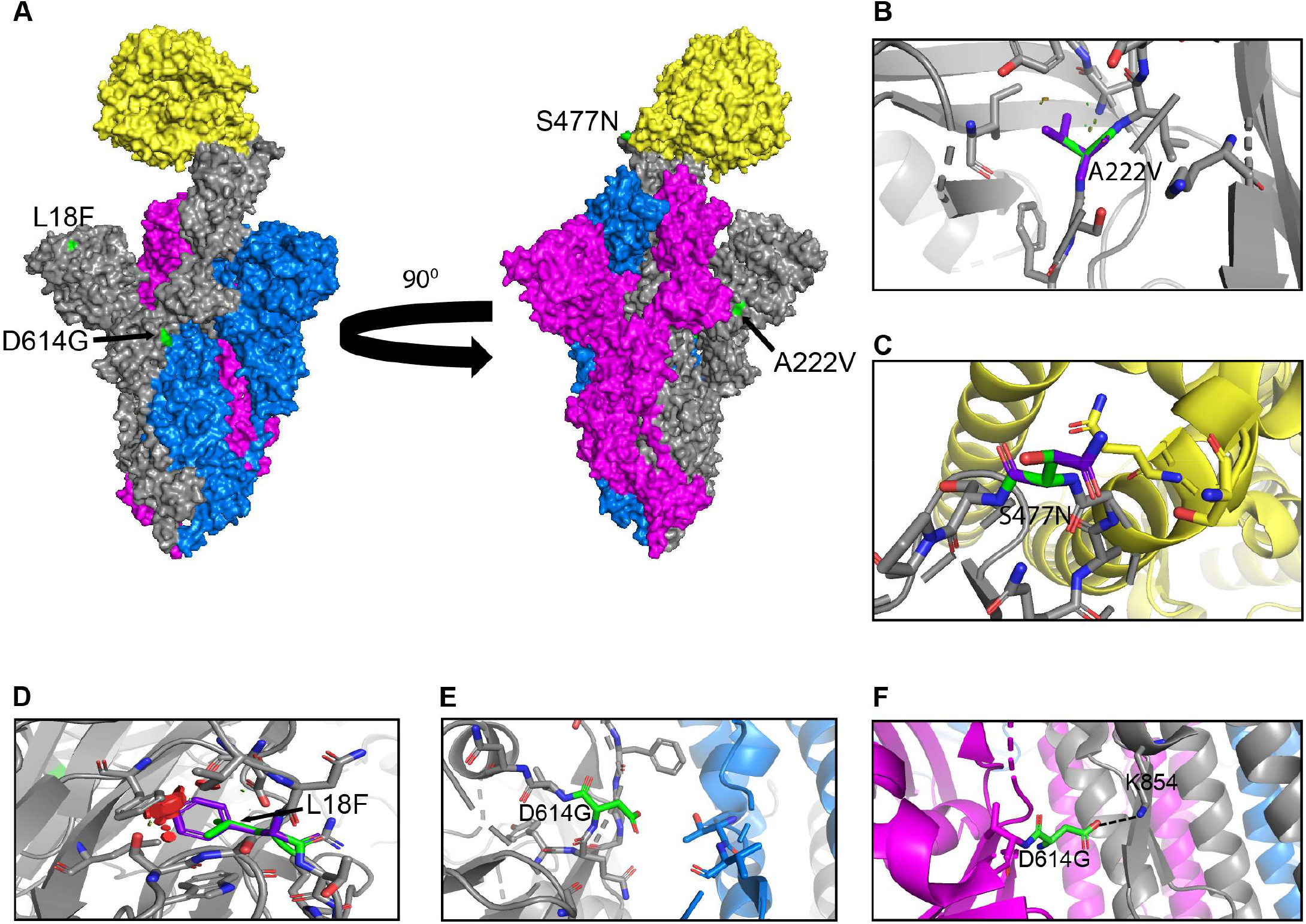
Structural rendering of the most frequent mutations in the S protein. A) Surface representation of hACE2 (yellow) in complex with S protein trimers illustrated in grey, blue, and magenta. Interactions of high frequency mutations are presented as follows: B) A222V, C) S477N, D) L18F, E) D614G in the open conformation and F) D614G in the closed conformation. Reference sequence residues are illustrated in green, and the mutated amino acid is represented in purple. The red markers illustrate the steric clash when the mutations are inserted into the structure. Graphs were generated using PyMOL.

### The emergence of the B.1.1.7 variant in the UK

A new variant was discovered in late 2020 in the UK that displayed increased affinity to hACE2, virulence and Nabs escape capabilities (**Fig. 4**) (24,25,30,31). Here we further investigated the B.1.1.7 variant by looking at S protein mutations of this variant in complex with Nabs and hACE2. We mapped the localization of mutations with available Cryo-EM structures of the S protein and assessed the frequency of the variant by interrogating the GISAID database. There are nine mutations in the S protein out of the total 24 mutations in the B.1.1.7 SARS-CoV-2 genome (**Fig. 5A & 5E)**. Mutations in the S protein of the B.1.1.7 variant, except D614G, emerged in October of 2020 and reached a worldwide frequency of 70% to 80% in April 2021 (**Fig. 4, Fig. 5C, Table 2)**. N501Y, found in the RBD, can interact with K353 in hACE2 (**Fig. 5B & 5D, Fig. 8C)**. The N501Y mutation is associated with an increased affinity to hACE2, along with an increase in infectivity and virulence (**Table 2). Figure 5G** illustrates the whole genome of SARS-CoV-2 with all nucleotide substitutions of the B.1.1.7 variant. Substitutions C913T, C5986T, C14676T, C15279T, C16176T in ORF1ab, and T26801C in M protein are all synonymous mutations. Also, the C27972T mutation has a frequency of 75% and produces a premature stop codon in NS8 that inactivates the protein (Q27stop) without obvious consequences (**Fig. 5E, Table 2)** (32). These results allow us to better understand the frequencies, localization, and interactions of mutations in the S protein of the B.1.1.7 variant. For viruses, both synonymous and non-synonymous mutations are of importance. Synonymous mutations that don’t change the amino acid sequence of proteins can still exercise very important functions. They can play a key role in docking sites for RNA binding proteins, transcription factors and primers, or partake in important secondary structures of the viral RNA like functional loops and folds.

**Figure 4:**
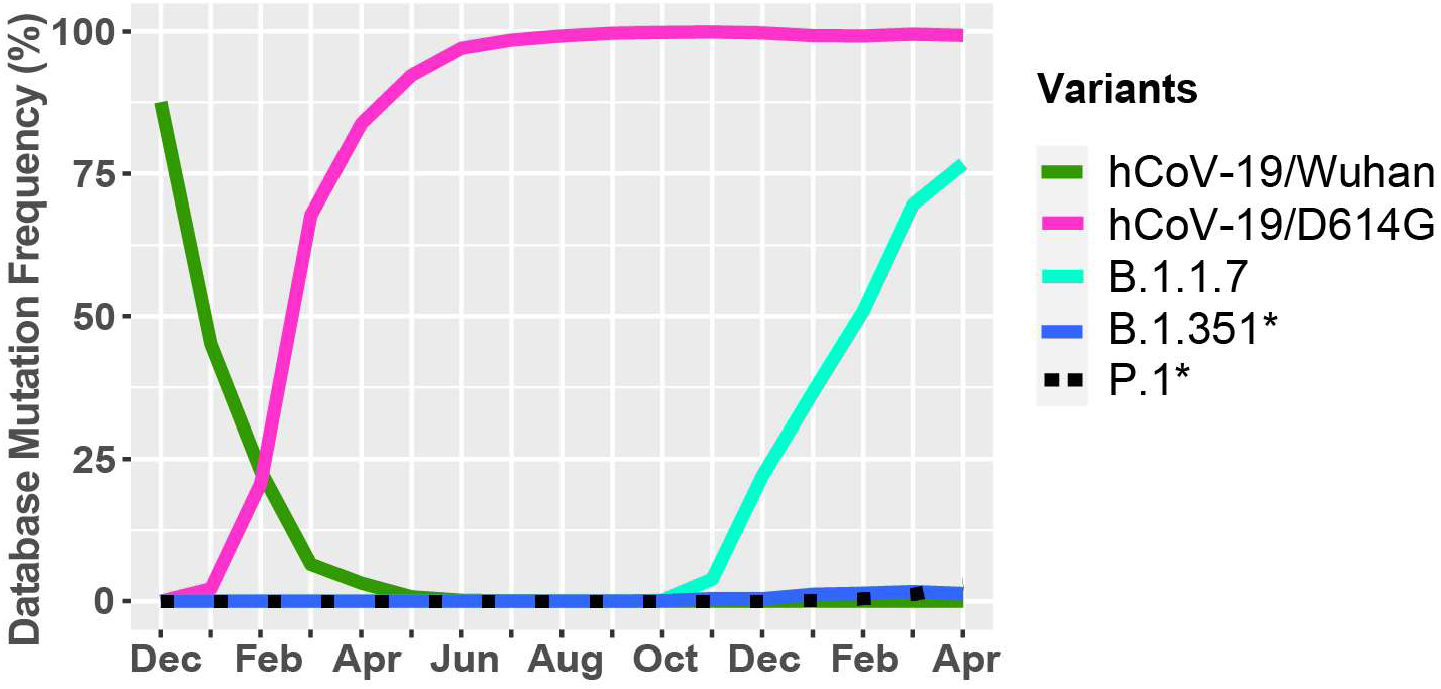
Worldwide frequency of B.1.1.7, B.1.351, P.1 and D614G variants. Database variants frequency were analyzed from December, 2019 to April 30^th^, 2021. hCoV-19/Wuhan is the original reference strain and hCoV-19/D614G is the current reference strain containing the D614G mutation in the S protein. B.1.1.7, B.1.353, and P.1 represent the UK, South African, and Brazilian variants. * overlapping curves.

**Figure 5:**
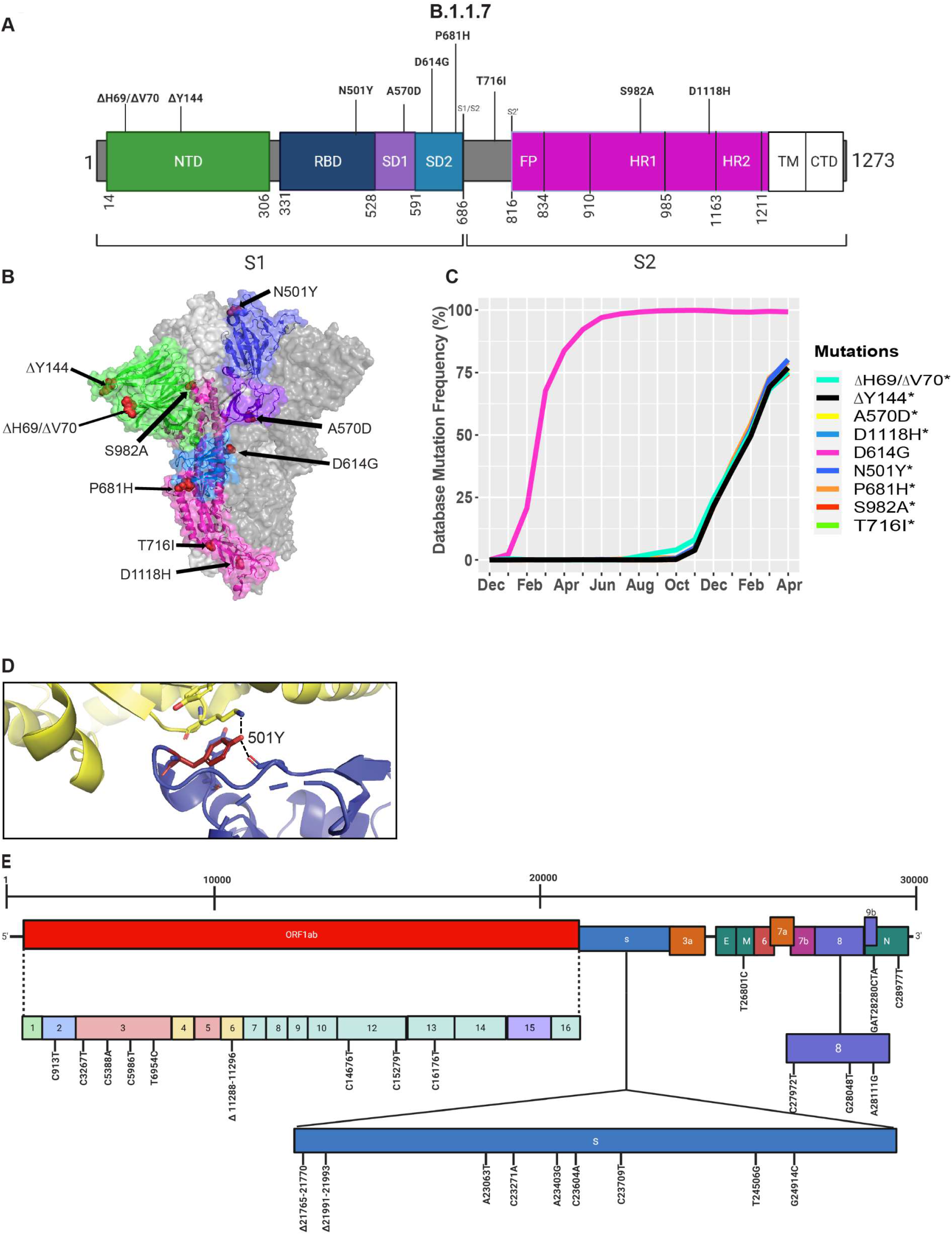
Analysis of mutations in the B.1.1.7 variant. A) Mutation map of the spike protein of B.1.1.7. B) Structural representation of spike with ACE2. B.1.1.7 S protein mutations are presented in in red. NTD (green), RBD (blue), SD1 (purple), SD2 (light blue), and S2 (magenta) are illustrated. The other S protein monomers are displayed in grey and white. C) Frequency of the mutations in the S protein, B.1.1.7 variant from December, 2019 to April 30^th^, 2021. D) Interaction of the N501 Y (red) mutation in the RBD (blue) of S protein with hACE2 (yellow). The dash lines indicate interactions with adjacent residues. E) Genome of the SARS-CoV-2 B.1.1.7 variant with identified nucleotide substitutions and deletions. Graphs were generated using Biorender, PyMOL, and RStudio. *overlapping curves.

### The emergence of the B.1.351 variant in South Africa

During the spread of the B.1.1.7 variant in the UK, another variant was emerging in South Africa, known as B.1.351 (21,22). The GISAID database was used to identify mutations and Cryo-EM structures of the S protein to model the effects of point mutations (**Fig. 6**). Most of the mutations in the S protein of the B.1.351 variant are localized in the S1 subunit, with only A701V in the S2 subunit. Additionally, three mutations reside in the RBD, among which two of them are not found in the B.1.1.7 variant (K417N, E484K) (**Fig. 6A, 6B**). However, this variant contains D614G and N501Y, which are also seen in the B.1.1.7 variant (**Fig. 5A)**. Furthermore, many of the mutations found in the B.1.351 variant have worldwide frequencies lower than 5%. The exception is D614G, L18F and N501Y (**Fig. 6C, Table 3)**. In comparison to B.1.1.7 and P.1 variants, the B.1.351 variant has not reached a frequency higher than 2% as of April 2021 (**Fig. 4**). By using the mutagenesis tool of PyMOL, we modeled the S protein in complex with hACE2, and with C103 and C121 Nabs, which are human recombinant class I & II neutralizing antibodies, respectively (33). Our *in silico* mutagenesis modelling predicts that B.1.352 mutations in the RBD favor a loss of interactions with C102 and C121 Nabs. Diminished interactions between RBD and C102 are predicted when the K417 is mutated to Asn producing Nabs escape capabilities (**Fig. 6D, 8A)**. Diminished interactions with RBD are also predicted with C121 when the E484K mutation is present (**Fig. 6E, 8B)**. As previously mentioned, N501Y is also found in B.1.351 and our modeling predicts that it will have similar effects as those observed with the B.1.1.7 variant (**Fig. 6F**). **Figure 6G** illustrates the position of the nucleotide substitutions of the B.1.351 variant.

**Figure 6:**
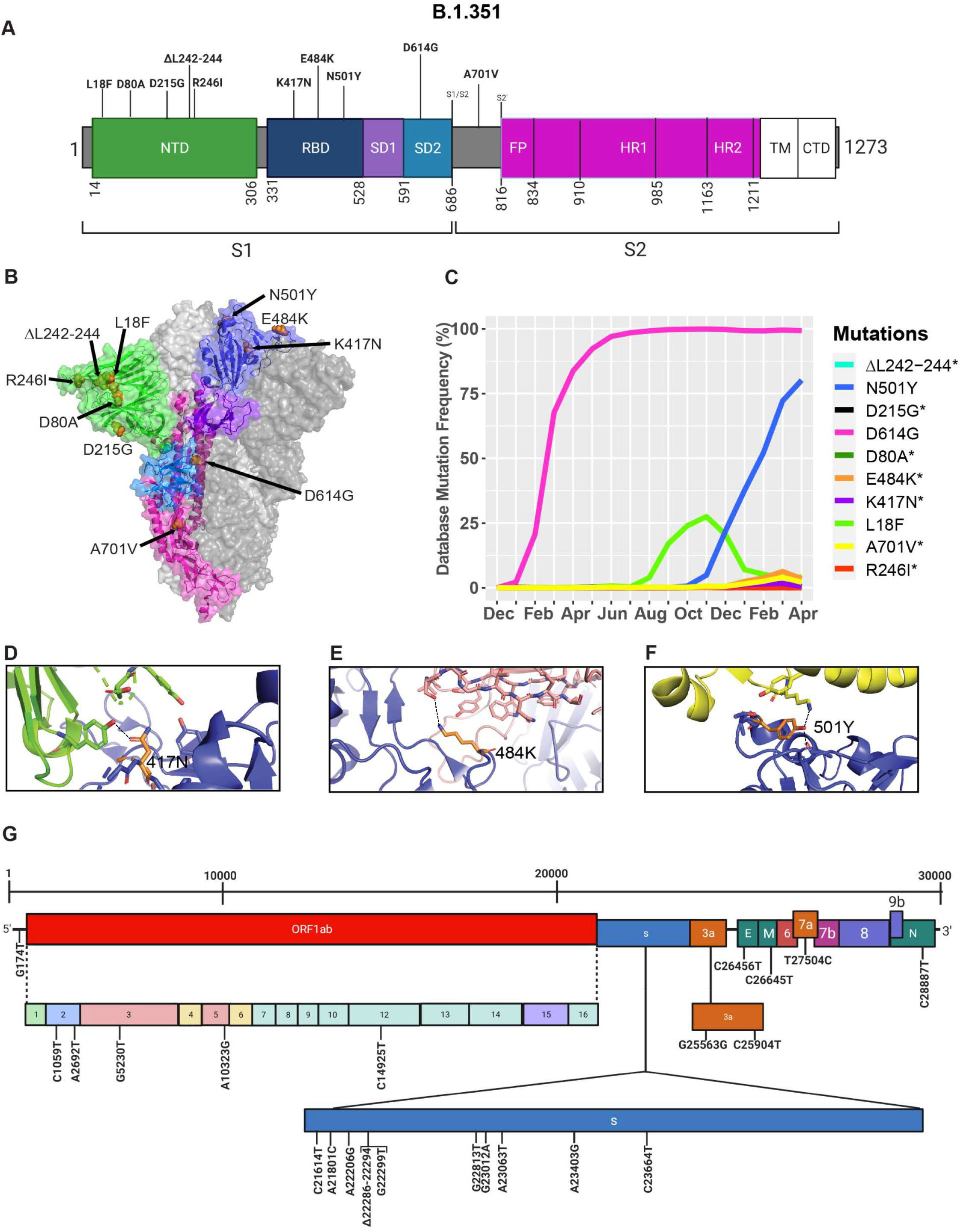
Analysis of mutations in the B.1.351 variant. A) Mutation map of the spike protein of B.1.351. B) Structural representation of spike with ACE2. B.1.352 S protein mutations are presented in orange. NTD (green), RBD (blue), SD1 (purple), SD2 (light blue), and S2 (magenta) are illustrated. The other S protein monomers are illustrated in grey and white. C) Frequency of the mutations in the S protein B.1.351 variant from December, 2019 to April 30^th^, 2021. Interaction of D) 417N with C102 Nab (green), E) 484K with C121 Nab (light pink), and F) 501Y with hACE2 (yellow). The mutant residues are illustrated in orange, and the dashed lines represent interactions with adjacent residues. G) Genome of the SARS-CoV-2 B.1.351 variant with identified nucleotide substitution s or deletions. Graphs were generated using Biorender, PyMOL, and RStudio. *overlapping curves.

**Table 3:**
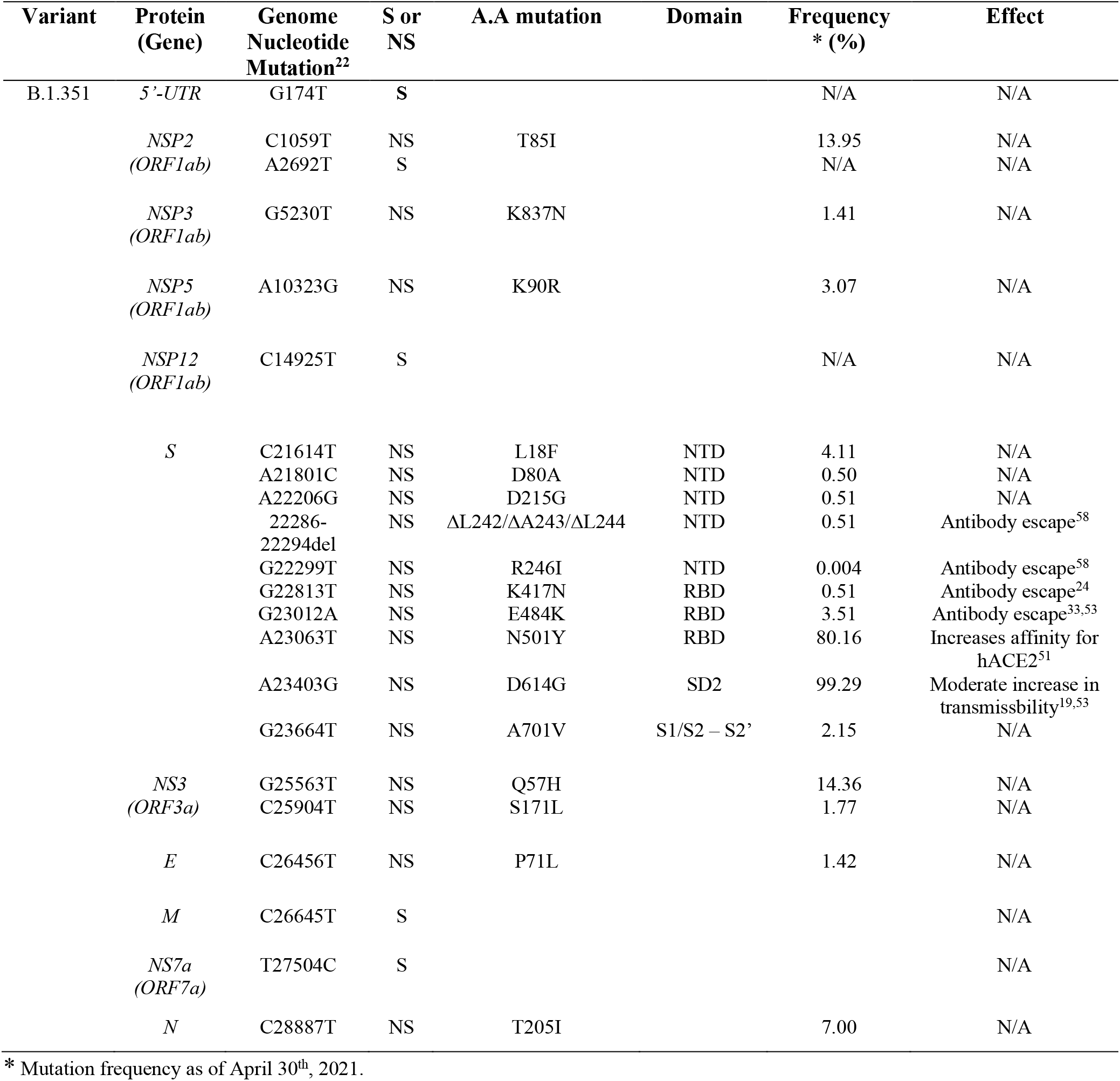
Synonymous, non-synonymous mutations and deletions in the B.1.351 variant

### The emergence of the P.1 variant in Brazil

Similar to the B.1.351 variant, the P.1 variant harbours the N501Y and E484K mutations, but position 417 of the S protein displays a threonine (T) instead of a lysine (K) residue (23) (**Fig. 7**). Similar to the UK and South African variants, we present in **Figure 7C** mutations in the S protein that are characteristic of the P.1 variant and their frequencies from December 2019 to April 30^th^ 2021. The P.1 variant harbours amino acid substitutions L18F, T20N, P26S, D138Y, and R190S in the NTD of the S protein. H655Y and T1027I, V1176F are in the subdomain 2 (SD2) of S1 and in the S2 subunit, respectively (**Fig. 7A, 7B, Table 4)**. The V1176F mutation is not shown because the structure is unresolved in this region. Overall, P.1-specific mutations have currently worldwide frequencies less than 5% (**Fig. 7C, Table 4)**. D614G, L18F and N501Y are not specific to P1. Similar to B.1.351, the P.1 variant has a low frequency representing less than 5% of global variants as of April, 2021 (**Fig. 4**). Here, we also modeled P.1 mutations to investigate interaction alterations with known recombinant neutralizing antibodies (33). The K417T mutation reduces interactions with neighboring residues in the C102 Nab and therefore the model predicts, as with K417N in B.1.1.7, an increased ability to escape neutralization (**Fig. 7D**). Also, the P.1 variant has E484K and N501Y mutations in the RBD. Modelling predicts they will have the same effect reported for the B.1.1.7 and B.1.351 variants (**Fig. 7E, 7F)**. In **Figure 7G**, we indicate the positions of synonymous, non-synonymous, and deletions in the SARS-CoV-2 P.1 genome.

**Figure 7:**
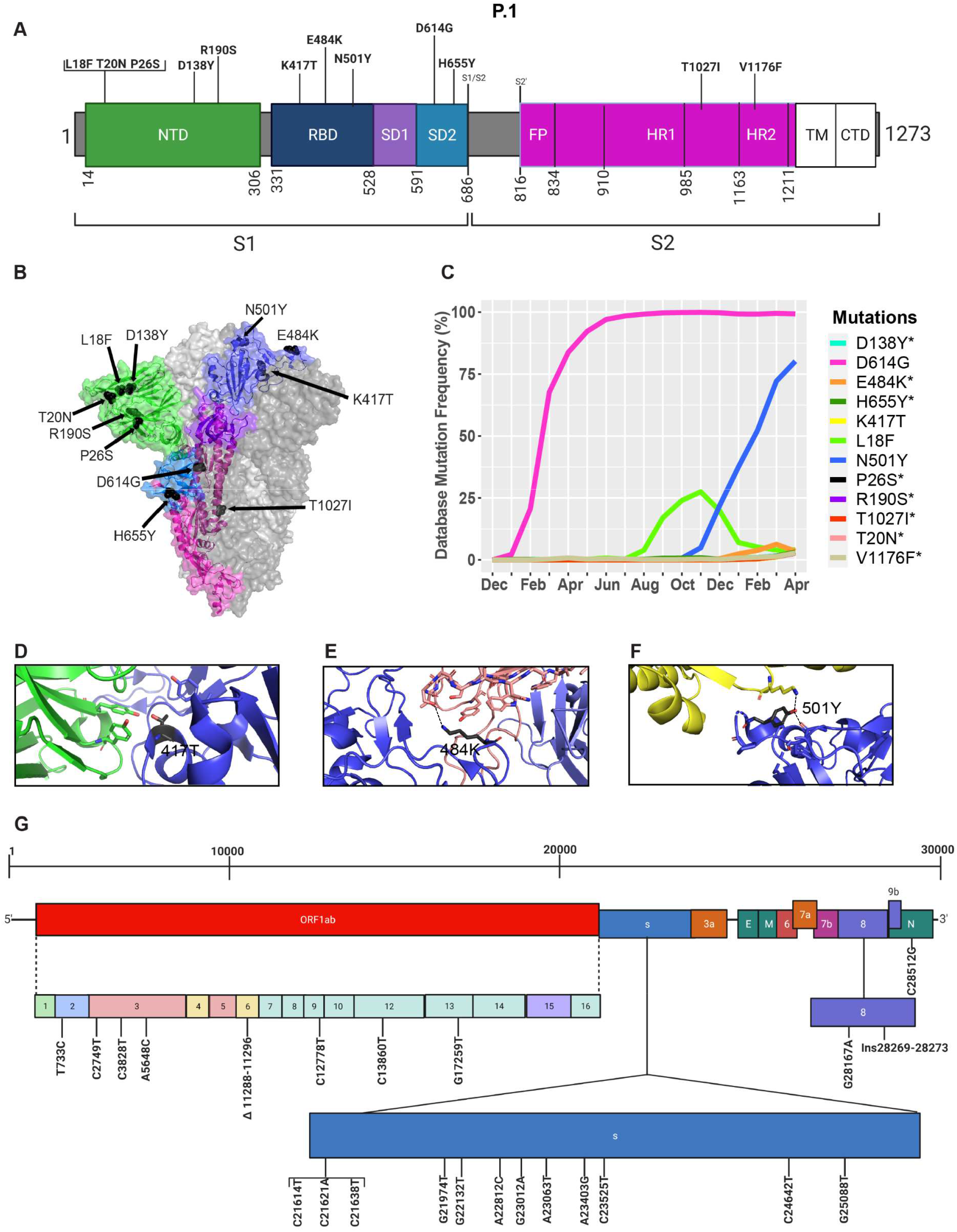
Analysis of mutations in the P.1 variant. A) Mutation map of the spike protein of P.1. B) Structural representation of spike with ACE2. P.1 S protein mutations are presented in black. NTD (green), RBD (blue), SD1 (purple), SD2 (light blue), and S2 (magenta) are illustrated. The other S protein monomers are illustrated in grey and white. C) Frequency of P.1 variant S protein mutations from December 2019 to April 30^th^ 2021. Interaction of D) 417T with C102 Nab (green), E) 484K with C121 Nab (light pink), and F) 501Y with hACE2 (yellow). The mutations are coloured in black and interaction with adjacent residues are demonstrated by dashed lines. G) Genome of the SARS-CoV-2 P.1 variant with identified nucleotides substitution, deletions, and insertions. Figures were generated using Biorender, PyMOL and RStudio. *overlapping curves.

**Table 4:** Synonymous, non-synonymous and deletions in the P.1 variant.

| Variant | Protein<br>(Gene) | Genome<br>Nucleotide<br>Mutation<sup>23</sup> | S or<br>NS | A.A<br>mutation | Domain | Frequency*<br>(%) | Effect |
| --- | --- | --- | --- | --- | --- | --- | --- |
| P.1 | *NSP2*<br>(*ORF1ab*) | T733C | S |  |  | N/A | N/A |
|  |  | C2749T | S |  |  | N/A | N/A |
|  | *NSP3*<br>(*ORF1ab*) | C3828T | NS | S370L |  | 3.09 | N/A |
|  |  | A5648C | NS | K977Q |  | 3.00 | N/A |
|  | *NSP6*<br>(*ORF1ab*) | 11288-11296del | NS | ΔS106/<br>ΔG107/ΔF<br>108 |  | 87.61 | N/A |
|  | *NSP9*<br>(*ORF1ab*) | C12778T | S |  |  | N/A | N/A |
|  | *NSP12*<br>(*ORF1ab*) | C13860T | S |  |  | N/A | N/A |
|  | *NSP13*<br>(*ORF1ab*) | G17259T | NS | E341D |  | 2.97 | N/A |
|  | *S* | C21614T | NS | L18F | NTD | 4.11 | N/A |
|  |  | C21621A | NS | T20N | NTD | 2.77 | N/A |
|  |  | C21638T | NS | P26S | NTD | 3.01 | N/A |
|  |  | G21974T | NS | D138Y | NTD | 2.95 | N/A |
|  |  | G22132T | NS | R190S | NTD | 2.71 | N/A |
|  |  | A22812C | NS | K417T | RBD | 2.88 | Antibody escape<sup>39</sup> |
|  |  | G23012A | NS | E484K | RBD | 3.51 | Antibody escape<sup>33,53</sup> |
|  |  | A23063T | NS | N501Y | RBD | 80.16 | Increase affinity for<br>hACE2<sup>53</sup> |
|  |  | A23403G | NS | D614G | SD2 | 99.29 | Moderate increase in<br>transmissibility<sup>19,53</sup> |
|  |  | C23525T | NS | H655Y | SD2 | 3.16 | N/A |
|  |  | C24642T | NS | T1027I | S2 | 2.79 | N/A |
|  |  | G25088T | NS | V1176F | S2 | 2.79 | N/A |
|  | *NS8*<br>(*ORF8*) | G28167A | NS | E92K |  | 2.96 | N/A |
|  |  | Ins28269-28273 | S |  |  | N/A | N/A |
|  | *N* | C28512G | NS | P80R |  | 2.94 | N/A |

\* Mutation frequency as of April 30<sup>th</sup>, 2021.

**Figure 8:**
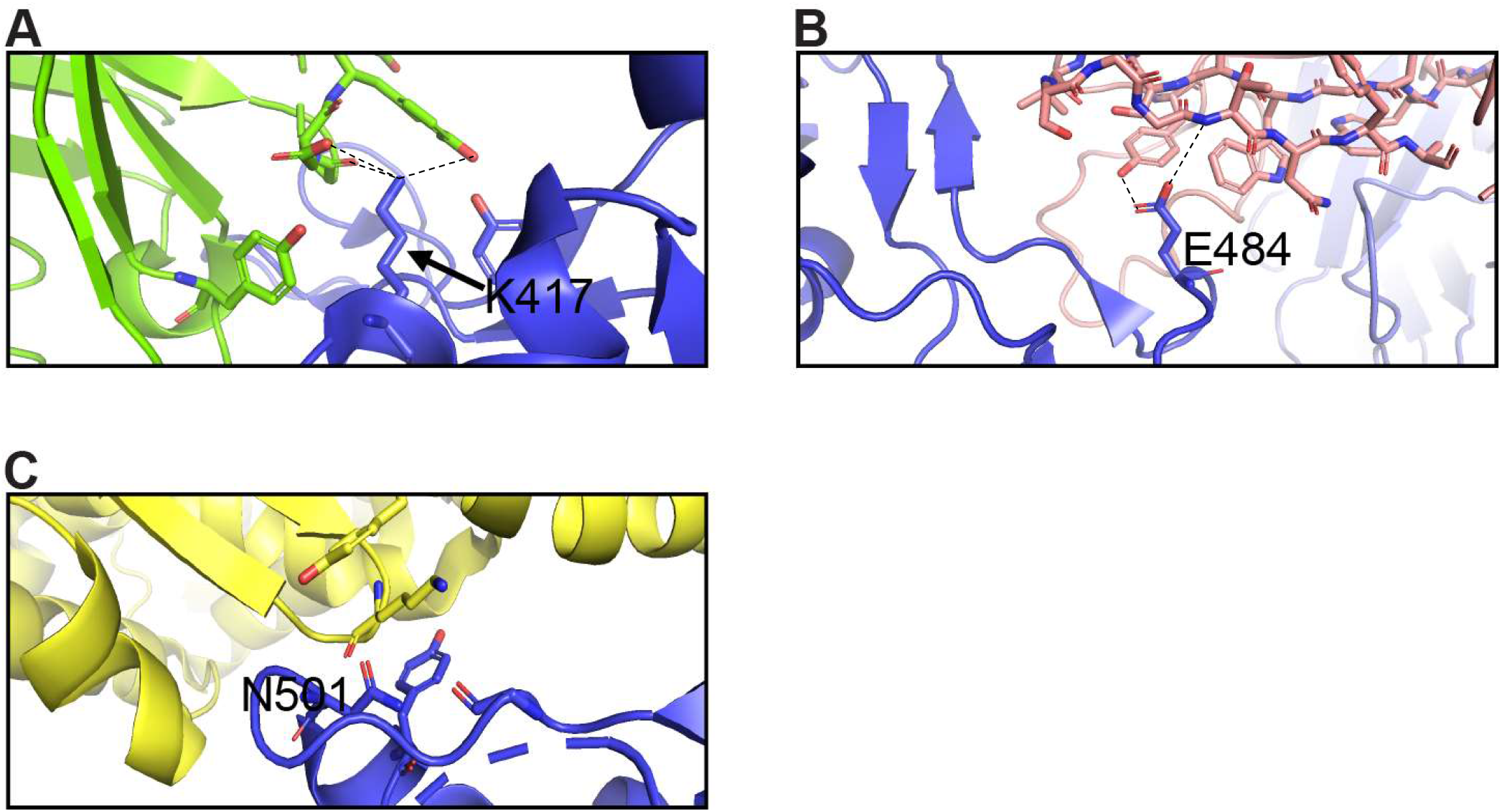
Interactions of K417, E484, and N501 of the S protein with neutralizing antibodies and hACE2. A) Interaction of K417 (blue) with C102 Nab (green) residues. B) Interaction of E484 (blue) with C121 Nab (light pink). C) Interaction of N501 (blue) with hACE2 (yellow) Dashes lines indicate interactions between residues. The graphs were generated using PyMOL.

## DISCUSSION

The emergence of new genetic variants that are more transmissible, virulent and resistant to antibody neutralization have highlighted the importance of studying the function of mutations in the viral genome. The number of sequenced viral genomes uploaded to the GISAID database grew rapidly from 131,417 at the end of September, 2020 to 451,913 by January 30^th^, 2021 (28). GISAID is a formidable tool for tracking the emergence of mutations, identifying the geographic region where it emerged, and tracking its spread around the globe. Given the risk mutations and new variants pose for neutralizing antibody therapy and vaccines for SARS-CoV-2, it is crucial to continuously monitor the susceptibility of these variants to neutralization by humoral and cellular immune responses either induced through natural exposure to the reference strain or induced by vaccination (24,25,33). Recent reports on the efficacy of the various vaccines have shown that these are variable and often diminished against variants (**Tables 5 and 6**). Most available vaccines generally remain near fully efficacious against the B.1.1.7 variant (31,34,35). However, the Oxford-AstraZeneca vaccine has displayed compromised efficacy against the B.1.351 variant with only 10% vaccine efficacy (36). Preliminary data with the Pfizer-BioNTech and Moderna mRNA vaccines also show a reduction in efficacy against B.1.351 (35,37,38). Furthermore, antibodies induced by the Pfizer-BioNTech and Moderna vaccines appear to display a 6.7-fold, and 4.5-fold decrease in neutralization efficacy against the P.1 variant (**Table 6)** (39). The recently approved Johnson & Johnson adenovirus-based vaccine only requires a single dose, in comparison to two for the mRNA vaccines and Oxford-AstraZeneca vaccine, and has an efficacy of 66% against the original Wuhan reference strain, 52.0 to 64.0% against the B.1.351 variant and 66% against the P.1 variant (43). The Novavax vaccine also shows reduced efficacy against the variants with 86% and 51% against the B.1.1.7 and P.1 variants, respectively. Nevertheless, humoral responses are only one component of the adaptive immune response. T cell responses have not been studied in detail against these variants at this time and still provide robust contribution to overall protection against severe COVID-19 disease.

**Table 5:** Efficacy of Vaccines Against SARS-CoV-2 Variants

| **Vaccine** | **hCoV-19/Wuhan/WIV-4/2019** | **B.1.1.7** | **B.1.351** | **P.1** |
| --- | --- | --- | --- | --- |
| Pfizer-BioNTech | 95.0%<sup>40</sup> | 95.3%<sup>31,37</sup> | 75.0%<sup>37</sup> | N/A |
| Moderna | 94.1%<sup>41</sup> | N/A | N/A | N/A |
| Oxford-AstraZeneca | 70.4%<sup>42</sup> | 70.4%<sup>34</sup> | 10%<sup>36</sup> | N/A |
| Johnson & Johnson | 66%<sup>43</sup> | N/A | 52.0% and 64.0%<br>(14 and 28 days)<sup>43</sup> | 66%<sup>43</sup> |
| Novavax | 96%<sup>44</sup> | 86%<sup>44</sup> | 51%<sup>44</sup> |  |

**Table 6:** Resistance of SARS-CoV-2 Variants to Vaccine-Induced Antibody Neutralization

| **Vaccine** | **Fold Reduction in Neutralization** |  |  |
| --- | --- | --- | --- |
|  | **B.1.1.7** | **B.1.351** | **P.1** |
| Pfizer-BioNTech | <sup>a</sup>2.1<sup>39</sup> | <sup>a</sup>(34.5-42.4)<sup>39</sup> | <sup>a</sup>6.7<sup>39</sup> |
| Moderna | <sup>a</sup>2.3<sup>39</sup> | <sup>a</sup>(19.2-27.7)<sup>39</sup> | <sup>a</sup>4.5<sup>39</sup> |
| Oxford-AstraZeneca | <sup>b</sup>2.5<sup>45</sup> | <sup>b</sup>9.0<sup>46</sup> | <sup>b</sup>2.6<sup>47</sup> |
| Johnson & Johnson | N/A | N/A | N/A |
| Novavax | <sup>a</sup>(0.85 to 20)<sup>48</sup> | <sup>a</sup>13.1<sup>49</sup> | N/A |

<sup>a</sup>Pseudotyped virus neutralization assay.<sup>b</sup>Live virus neutralization assay.

Studying the effect of individual mutations in a viral protein may not always reflect their true impact on virus features if they appear in combination with other mutations. Epistasis is the combinatory effect of two or more mutations in a genome (50). Epistasis has previously been intensely studied in the surface protein hemagglutinin (HA) of influenza viruses and positive epistasis was identified in several regions of the HA receptor-binding domain (51). In relation to the S protein of SARS-CoV-2, epistatic mutations could, for instance, allow the S protein to adopt a specific conformation when all the mutations are present, thereby producing a unique folding of the protein and enabling new features. A recent study demonstrated the impact of antigenicity and infectivity of D614G SARS-CoV-2 variants with a combination of different mutations occurring in the S protein (19). The study shows that D614G alone only mildly increases infectivity, but in combination with different other mutations in the S protein, together these can either more intensely increase or decrease viral infectivity. Similar findings have been reported regarding sensitivity to Nabs. D614G alone has undetectable effects on Nabs escape. However, the combination of D614G with other mutations in S can enable Nabs escape (19). This data suggests that the continuous emergence of epistatic mutations in SARS-CoV-2 will likely be involved in further altering the properties of the virus, including transmissibility, pathogenicity, stability and Nab resistance.

Our analyses have highlighted that several new fast-spreading mutations had frequency trajectories that eventually plunged or stabilized at low frequencies. Only the D614G in the S protein and the P323L in the RdRp have maintained their presence in the consensus sequence (**Fig. 1 and 2**). Analyses of the GISAID database also reveal which countries upload the most sequences to the database and are therefore carrying out the most testing and sequencing. The global frequency of mutations and variants in the database is therefore biased to represent the genetic landscape of the virus in the countries doing the most testing, sequencing, and data sharing. Emergence of new variants may therefore go undetected until they leave their point of origin and enter countries with high testing and sequencing rates. This delayed notification constitutes a major obstacle in identifying and preventing the spread of nefarious variants that are potentially resistant to current vaccines and neutralizing antibody therapy. Another caveat in analyzing sequences in the GISAID database is that consensus sequences are uploaded, but subsequences and quasispecies are generally not included. Therefore, it is possible that mutations in sub-variants may be missed that could contribute to virus replication, pathogenicity, and spread (52). A global approach to analyzing both transmitted variants and non-transmitted sub-variants and quasispecies could provide a better understanding of the effects of SARS-CoV-2 mutations.

In conclusion, our metadata analysis of emerging mutations has highlighted the natural fluctuations in mutation prevalence. We also illustrate how mutations sometimes need to co-emerge in order to create a favorable outcome for virus propagation. Tracking mutations and the evolution of the SARS-CoV-2 genome is critical for the development and deployment of effective treatments and vaccines. Thus, it is the responsibility of all countries and governing jurisdictions to increase testing and sequencing, and uploading SARS-CoV-2 genomes to open access databases in real-time. This will result in more accurate information to inform policy and decision makers about interventions required to blunt the global transmission of the virus and ensure that vaccines remain effective against all circulating variants.

## Data Availability

All sequence data analyzed is freely available on GISAID:

https://www.gisaid.org

## ACKNOWLEDGEMENTS

The authors wish to thank Dr. Sean Li at Health Canada for helpful comments on our manuscript. M.-A.L. holds a Canada Research Chair in Molecular Virology and Intrinsic Immunity. This study was supported by a COVID-19 Rapid Response grant to M-A Langlois by the Canadian Institute of Health Research (CIHR) and by a grant supplement by the Canadian Immunity Task Force (CITF).

## CONFLICTS OF INTERESTS

The authors declare no competing interests.

## Notes

### Competing Interest Statement

The authors have declared no competing interest.

### Author Declarations

This is metadata analysis of public sequences, IRB oversight was not required.

